# Progressivity of out-of-pocket costs under Australia’s universal health care system: a national linked data study

**DOI:** 10.1101/2022.05.29.22275749

**Authors:** HD Law, D Marasinghe, D Butler, J Welsh, E Lancsar, E Banks, N Biddle, R Korda

## Abstract

**Background:** In line with affordability and equity principles, Medicare—Australia’s universal health care program—has measures to contain out-of-pocket costs (OOPC), particularly for lower income households. This study examined the distribution of OOPC for Medicare-subsidised out-of-hospital services and prescription medicines in Australian households, according to their ability to pay.

**Methods:** OOPC for out-of-hospital services and medicines in 2017-18 were estimated for each household, using 2016 Australian Census data linked to Medicare Benefits Schedule (MBS) and Pharmaceutical Benefit Scheme (PBS) claims. We derived household disposable income by combining income information from the Census linked to income tax and social security data. We quantified OOPC as a proportion of equivalised household disposable income and calculated Kakwani progressivity indices (K).

**Results:** Using data from 85% (n=6,830,365) of all Census private households, OOPC as a percentage of equivalised household disposable income decreased from 1.16% in the poorest decile to 0.63% in the richest decile for MBS services, and from 1.35% to 0.35% for PBS medicines. The regressive trend was less pronounced for MBS services (*K* = −0.06), with percentage OOPC relatively stable between the 2^nd^ and 9^th^ income deciles; while percentage OOPC decreased substantially with increasing income for PBS medicines (*K* = −0.24).

**Conclusion:** OOPC for out-of-hospital Medicare services were mildly regressive while those for prescription medicines were distinctly regressive. Actions to reduce inequity in OOPC, particularly for medicines should be considered.

## INTRODUCTION

The goal of universal health care is to ensure people have access to essential health services without financial hardship.^1^ Most universal health care systems rely on patients paying some of the cost out of their own pockets. The affordability of these costs is an increasing global concern, both in absolute terms and the extent to which they are distributed equitably according to ability to pay.^2, 3^

Australia has a mixed public and private system. Medicare, Australia’s universal public health insurance scheme, provides free public hospital care. The scheme also subsidises private hospital care and a range of services provided out-of-hospital and prescription medicines. In 2019-20, Australians spent an estimated $29.8 billion out-of-pocket on health,^4^ including over $6 billion on Medicare-subsidised out-of-hospital services^5^ and prescription medicines.^6, 7^ Out-of-pocket costs for Medicare-subsidised out-of-hospital services and medicines cannot be claimed through private health insurance.

The Medicare Benefits Schedule (MBS) lists the services covered by Medicare, including those provided by general (primary care) practitioners and specialists. The MBS lists a price signal and the benefit amount (rebate) that it will provide. However, doctors are free to charge any price. Therefore, any excess fee charged above the benefit results in an out-of-pocket cost to the patient. Prescription medicines subsidised by Medicare are listed on the Pharmaceutical Benefits Scheme (PBS), where patients pay out-of-pocket up to a maximum amount (co-payment) per medicine.

Medicare has measures to help contain high out-of-pocket costs, particularly for lower income households. First, doctors can choose to bill Medicare directly (bulk-bill) and accept the benefit as full payment, making the service free to the patient.^8^ Medicare also provides incentive payments to doctors for bulk-billing concession card holders (available to pensioners and those receiving certain social security benefits, among other eligibility criteria).^9^

Second, there are safety net schemes. Under separate schemes, out-of-pocket costs for out-of-hospital MBS services and for PBS medicines are accumulated in the calendar year. Once the respective thresholds are reached, the Medicare Safety Net provides increased benefits to patients for subsequent services, while the PBS Safety Net acts to reduce the maximum co-payment for subsequent prescriptions; in both cases reducing the out-of-pocket costs incurred. Patients with concession cards are eligible for lower safety net thresholds. Families can also combine costs in order to reach the thresholds sooner.^8, 10^

These measures to contain out-of-pocket costs under Medicare are in line with affordability and equity principles of universal health care. However, the extent to which these costs are progressive (richer households contribute proportionally more of their income), proportional (all households contribute the same proportion of their income) or regressive (richer households contribute proportionally less of their income) is unclear.

A recent international report indicated that overall out-of-pocket costs in Australia are regressive.^3^ However, this was based on relatively small household surveys and included all health-related expenses including services not covered by Medicare (e.g. non-prescription medicines, dental services and most allied health). The aim of this study was to quantify out-of-pocket costs as a proportion of household disposable income for Medicare-subsidised out-of-hospital services and medicines in Australia, using linked administrative MBS, PBS and income data.

## METHODS

Briefly, we used Australian 2016 census data linked to Medicare claims data (to quantify out-of-pocket costs) and income tax and social security data (to estimate household disposable income). We then quantified out-of-pocket cost as a proportion of equivalised household disposable income at each income decile, and calculated Kakwani progressivity indices.

### Data sources and study population

We used de-identified 2016 Census of Population and Housing data linked individually to the Medicare Consumer Directory (MCD) (2006-2018), MBS data (2017-18), PBS data (2017-18), Personal Income Tax (PIT) data (2016-17), DOMINO Centrelink Administrative data (2016-17) and Death Registrations (2016-17), compiled through the Multi-Agency Data Integration Project (MADIP). Data were linked by the Australian Bureau of Statistics via the Person Linkage Spine, a person-level identification key that broadly covers the resident population of Australia from 2006.^11^

The scope of the 2016 Census was usual residents of Australia on 9 August 2016. The 2016 Census had a dwelling response rate of 95% and person response rate of 95%.^12^ The eligible population for this study was all private households in the 2016 Census that linked to the MCD. The proportion of individuals living in private households in the 2016 Census who linked to the MCD via the Person Linkage Spine was 93%.

We estimated income by combining income information from the Census, PIT and DOMINO. PIT data contain income amounts for those who completed a tax return in the financial year. DOMINO data contain the amounts of social security payments paid to those who were eligible. We used MBS and PBS claims data to calculate out-of-pocket costs for each individual. MBS data contain the fees charged and benefits paid for MBS-listed services, while PBS data contain patient contribution amounts for PBS-listed medicines.

The study sample comprised all occupied private households in the 2016 Census where every member was enrolled on the MCD by 1 July 2017. While income Is usually received by individuals, it is assumed to be shared between members of a household; therefore, the unit of analysis in this study was the household. We excluded households where at least one member was absent on Census night (no Census data) or without at least one adult. We also excluded households where at least one adult member: died between 9 August 2016 and 30 June 2017, did not have an income estimate, or was top-coded in PIT (total income ≥ $5 million). If a child member died between 9 August 2016 and 30 June 2017, the child was removed from the household and household size reduced accordingly. Supplementary Figure 1 shows a flow chart of sample selection.

### Study variables

#### Equivalised household disposable income

We estimated the equivalised household disposable income in order to standardise for variations in household size and composition, while taking into account the sharing of income within households. Firstly, we estimated individual disposable (or post-tax) income in the 2016-17 financial year using a combination of income information from the PIT, DOMINO and Census. We then summed individual disposable incomes across household members. We adjusted household income using the ‘modified OECD’ equivalence scale, which assumes the second and subsequent adult in the household costs 0.5 times the amount of the first adult, and children cost 0.3 times the amount of the first adult.^13^

Disposable income for children (<15 years) was zero. For adults, personal disposable income was estimated using the following formulae and hierarchy:

1. For adults who linked to PIT:

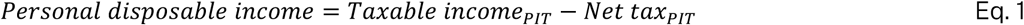 For the subset of adults aged ≥60 and not in the labour force, we added the amount of Census income in excess of PIT total income to disposable income calculated in Equation 1. This adjustment attempted to capture tax-exempt superannuation:

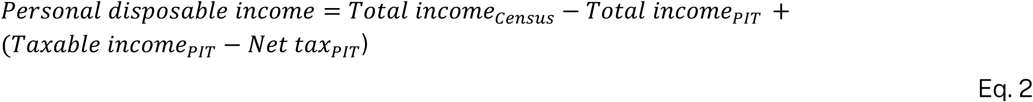
2. For adults who linked to DOMINO:

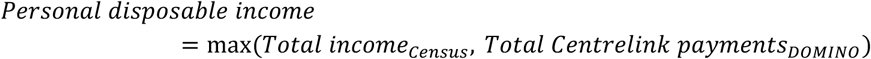
3. For adults without a link to either PIT or DOMINO:

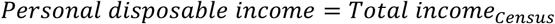

Income data are continuous in PIT and DOMINO but categorical in the Census.^a^ We predicted continuous Census income using an interval regression model^14^ with income categories as the dependent variable and the following independent variables from the Census: age, gender, Indigenous status, highest level of education, labour force status, occupation, industry of employment, severe disability and geographical area (Statistical Area Level 4).

We combined income data across collections given the limitations of each individual dataset. Chiefly, PIT is likely to have excluded those with lower incomes.^b^ Therefore, we estimated income for this group from Census or DOMINO. However, there are differences between these data: 1) Census is self-reported, while PIT and DOMINO are administrative, and 2) Census asks for total income, while PIT collects information on income within the tax system only.^c^ While we could have used Census as the only source of income data for those that did not have PIT data, empirically, those in this group tended to self-report a lower Census income in comparison to total Centrelink payments (Supplementary Figure 2). We therefore took the higher of these figures.

#### Out-of-pocket cost

The outcomes in this study were annual per household out-of-pocket costs, in the 2017-18 financial year for: out-of-hospital MBS services and PBS medicines, each as a proportion of equivalised household disposable income.

For MBS services, out-of-pocket cost per service was calculated as fee charged less benefit paid, in Australian dollars. These were then summed across all claims made in 2017-18 for each household. The cost of bulk-billed services was set to zero. For PBS medicines, the patient contribution amount was summed across all medicines bought in 2017-18 for each household. Individuals with no record in the MBS or PBS datasets had zero out-of-pocket cost.

In addition, we calculated the proportion of households in each income decile whose annual out-of-pocket costs reached the lower (concessional) safety net thresholds, thus potentially eligible for reduced out-of-pocket costs (Supplementary Table 1). It was not possible to determine concessional entitlements; therefore, by applying the lower concessional threshold, we over-estimated the number of households that were captured by the safety nets in actuality.

### Statistical analysis

We calculated the mean household out-of-pocket cost as a proportion of equivalised household disposable income for each income decile. Households in the first and second percentiles were excluded, such that the bottom decile contained 8% of households. This excluded very low income households who may be supported by high levels of wealth, consistent with ABS methods.^15^ In supplementary analyses, we stratified households by family type, excluding multiple-family households.

#### Concentration curves

We used the Kakwani index as a summary measure of equity in the distribution of out-of-pocket cost in relation to household income.^16^ The Kakwani index is defined as *K* = *C* – *G*, where *C* and *G* are the concentration indices for cost and income (also known as Gini coefficient) respectively.

The concentration curve plots the cumulative percentage of households ranked from poorest to richest on the *x* axis, and the cumulative percentage of income (Lorenz curve) or cost on the *y* axis. Therefore, the curves show, respectively, the income and cost share of any selected cumulative proportion of the population. If the cumulative share of income or cost is lower than the share of the population, the curve falls below the line of equality (where share of income or cost = share of population), and vice-versa. The extent to which the Lorenz curve lies outside (inside) the cost concentration curve represents the degree of regressivity (progressivity) of costs and is quantified by *K* (twice the area in between the Lorenz and cost curve). *K* lies between −2 and 1, and is positive if cost is progressive, negative if cost is regressive and zero if cost is proportional to income.

#### Sensitivity analyses

In sensitivity analyses, we re-calculated *K* in two ways that did not require combining income across different collections. First, we used income data exclusively from PIT, noting that this sample represented taxpaying households only. Secondly, we used income data from the Census only, noting that this was a total income measure inclusive of tax and other deductions therefore overestimated the disposable income of taxpayers.

## RESULTS

### Sample description

There were 8,281,631 private households in the 2016 Census, occupied by 20,825,664 residents. After excluding households that did not have an active enrolment on the MCD (and other exclusions, see Appendix Figure 1), our final sample consisted of 6,830,365 private households (82%) with 16,921,946 residents (81%).

Among those in our sample, 73% of adults had a PIT record and 40% had a DOMINO record in 2016-17 (Table 1). A very high percentage of individuals had at least one MBS claim (91%) and 71% had at least one PBS claim in 2017-18. Our sample was very similar to private households in the Census as a whole (Table 1), with one exception: our sample had a higher proportion of lone person households, likely due to our condition that all household members had to be linked to the MCD.

**Table 1.** Demographic and socioeconomic characteristics for individuals and private households in the Census vs. our study sample, 2016.

| Characteristic | 2016 Census private households | Sample |
| --- | --- | --- |
| *Individuals* |  |  |
| Total sample | 20,825,664 | 16,921,946 |
| **Sex** |  |  |
| Male | 49% | 49% |
| Female | 51% | 51% |
| **Indigenous status** |  |  |
| Non-Indigenous | 96% | 97% |
| Aboriginal and/or Torres Strait Islander | 3% | 2% |
| Unknown | 1% | 1% |
| **Age group** |  |  |
| 0-14 | 20% | 20% |
| 15-64 | 66% | 64% |
| 65-84 | 13% | 14% |
| 85+ | 2% | 2% |
| **Remoteness Area** |  |  |
| Major cities | 72% | 72% |
| Inner regional | 18% | 19% |
| Outer regional | 8% | 8% |
| Remote | 1% | 1% |
| Very remote | 1% | 0% |
| **Labour force status (adults)** |  |  |
| Employed | 61% | 61% |
| Unemployed | 4% | 4% |
| Not in labour force | 34% | 34% |
| Unknown | 1% | 1% |
| **Linked to Person Linkage Spine** |  |  |
| Yes | 95% | 100% |
| No | 5% | 0% |
| **Has PIT data (adults)** |  |  |
| Yes | 69% | 73% |
| No | 26% | 27% |
| Not linked to Spine | 5% | N/A |
| **Has DOMINO data (adults)** |  |  |
| Yes | 37% | 40% |
| No | 58% | 60% |
| Not linked to Spine | 5% | N/A |
| **Has out-of-hospital MBS record** |  |  |
| Yes (mean/median annual OOP*) | 85% (\$151/ \$21) | 91% (\$155/ \$26) |
| No | 10% | 9% |
| Not linked to Spine | 5% | N/A |
| **Has PBS record** |  |  |
| Yes (mean/median annual OOP*) | 65% (\$179/ \$89) | 71% (\$182/ \$93) |
| No | 30% | 29% |
| Not linked to Spine | 5% | N/A |
| **Mean/median Census total income based on midpoints** | **\$45,197/ \$37,700** | **\$46,452/ \$37,700** |
| *Households* |  |  |
| Total sample | 8,281,631 | 6,830,365 |
| **Mean/median household size** | **2.5/ 1.4** | **2.5/ 1.4** |
| **Household composition†** |  |  |
| Couple with no children | 27% | 27% |
| Couple with children | 32% | 31% |
| One parent family | 11% | 10% |
| Other family | 1% | 1% |
| Lone person household | 24% | 27% |
| Group household | 4% | 3% |
| **Mean/median Census equivalised total household income based on midpoints** | **\$54,532/ \$46,800** | **\$55,420/ \$46,800** |
| **Equivalised household disposable income ranges‡** |  |  |
| <\$21,572 | N/A | 10% |
| \$21,572–\$24,306 | N/A | 10% |
| \$24,306–\$29,803 | N/A | 10% |
| \$29,803–\$35,250 | N/A | 10% |
| \$35,250–\$41,008 | N/A | 10% |
| \$41,008–\$47,712 | N/A | 10% |
| \$47,712–\$55,580 | N/A | 10% |
| \$55,580–\$65,413 | N/A | 10% |
| \$65,413–\$82,339 | N/A | 10% |
| >\$82,339 | N/A | 10% |

Notes:
\* Mean/median OOP costs exclude those without any MBS/PBS record
† For multiple family households, the composition of the primary family is used.
‡ Derived from a combination of income data from Personal Income Tax, DOMINO Centrelink Data and Census for our study sample (see Methods).

### Out-of-pocket costs

In our sample, higher income was associated with lower average age of adults in the household, and volume of medicines and services accessed (Table 2). Higher income was also associated with a lower proportion of MBS services that were bulk-billed.

**Table 2.** Sample characteristics, service usage and annual out-of-pocket (OOP) costs for Medicare-subsidised services by income decile, Australian households, financial year 2017-18.

| Income decile | Sample |  | MBS services |  |  |  |  | PBS medicines |  |  |  |
| --- | --- | --- | --- | --- | --- | --- | --- | --- | --- | --- | --- |
| | Mean Household size | Mean Age of adults | Mean Number of services | Proportion Bulk-billed (%) | Mean OOP (SD) (\$) | Median OOP (Q1-Q3) (\$) | Mean OOP/ equivalised income (%) | Mean Number of supplies | Mean OOP (SD) (\$) | Median OOP (Q1-Q3) (\$) | Mean OOP/ equivalised income (%) |
| 1 | 2.5 | 48 | 37 | 0.91 | 189 (392) | 40 (0-231) | 1.16 | 29 | 221 (275) | 134 (38-324) | 1.35 |
| 2 | 1.7 | 65 | 41 | 0.92 | 164 (318) | 42 (0-210) | 0.70 | 48 | 268 (218) | 248 (99-396) | 1.15 |
| 3 | 2.5 | 53 | 46 | 0.91 | 211 (396) | 68 (0-276) | 0.79 | 45 | 290 (288) | 235 (77-416) | 1.08 |
| 4 | 2.6 | 52 | 44 | 0.89 | 251 (442) | 101 (0-334) | 0.78 | 41 | 315 (328) | 239 (82-437) | 0.98 |
| 5 | 2.6 | 49 | 42 | 0.87 | 294 (497) | 131 (0-393) | 0.77 | 36 | 327 (364) | 229 (74-446) | 0.86 |
| 6 | 2.7 | 46 | 39 | 0.86 | 335 (575) | 152 (0-438) | 0.76 | 29 | 337 (401) | 209 (63-457) | 0.76 |
| 7 | 2.7 | 44 | 39 | 0.84 | 392 (651) | 190 (27-507) | 0.76 | 27 | 353 (424) | 210 (63-477) | 0.69 |
| 8 | 2.6 | 44 | 36 | 0.82 | 439 (731) | 214 (35-561) | 0.73 | 24 | 351 (436) | 197 (56-474) | 0.59 |
| 9 | 2.5 | 43 | 35 | 0.79 | 525 (872) | 263 (54-655) | 0.72 | 22 | 350 (444) | 186 (52-474) | 0.48 |
| 10 | 2.4 | 46 | 34 | 0.74 | 697 (1,124) | 358 (90-862) | 0.63 | 22 | 371 (473) | 194 (51-506) | 0.34 |
| All | 2.5 | 49 | 39 | 0.86 | 353 (669) | 141 (0-439) | 0.77 | 32 | 320 (378) | 210 (64-432) | 0.82 |

Notes:

In absolute terms, household out-of-pocket costs for MBS services and PBS medicines in 2017-18 were right skewed, where the mean was always higher than the median at every income decile (Table 2). That is, most households had low or average costs while some households had very high costs. Therefore, we discuss average costs in terms of the median.

For MBS services, the median (and mean) out-of-pocket costs increased with equivalised household disposable income from $40 in the poorest decile to $358 in the richest decile (Table 2). For PBS medicines, median cost decreased with income from $248 in the second poorest decile to $194 in the richest decile, but the opposite was true for the mean (increased with income) (Table 2). This indicated an increasing right skew of PBS costs with increasing income i.e. as income increased, there were more and more households with very high PBS costs.

For both MBS and PBS, there was greater variation in costs as income increased, even as richer households tended to access fewer numbers of services and medicines (Table 2).

The above trends were reflected in the concentration indices (Table 3). The positive concentration index for MBS out-of-pocket costs (*C*_*MBS*_ = 0.25) indicated a ‘pro-rich’ distribution, where richer households paid an increasing share of total cost, from 4.4% in the poorest decile to 20.2% in the richest decile (Table 3). In contrast, the distribution of PBS out-of-pocket costs was only mildly pro-rich (*C*_PBS_ = 0.07). From the fourth income decile onwards, each decile paid about the same share (10-11%) of total cost.

**Table 3.** Cumulative distribution of out-of-pocket (OOP) costs for Medicare-subsidised services by equivalised household disposable income decile, Australian households, financial year 2017-18.

| Income decile | Households |  |  | Equivalised income |  | OOP costs |  |  |  |
| --- | --- | --- | --- | --- | --- | --- | --- | --- | --- |
|  | N | % | Cum. % | % | Cum. % | MBS services |  | PBS medicines |  |
|  |  |  |  |  |  | % | Cum. % | % | Cum. % |
| 1 | 546,434 | 8.2 | 8.2 | 2.8 | 2.8 | 4.4 | 4.4 | 5.6 | 5.6 |
| 2 | 683,034 | 10.2 | 18.4 | 4.8 | 7.6 | 4.7 | 9.1 | 8.5 | 14.2 |
| 3 | 683,035 | 10.2 | 28.6 | 5.6 | 13.2 | 6.1 | 15.2 | 9.2 | 23.4 |
| 4 | 683,037 | 10.2 | 38.8 | 6.7 | 19.9 | 7.3 | 22.5 | 10.0 | 33.4 |
| 5 | 683,036 | 10.2 | 49.0 | 7.9 | 27.8 | 8.5 | 31.0 | 10.4 | 43.9 |
| 6 | 683,037 | 10.2 | 59.2 | 9.2 | 37.0 | 9.7 | 40.6 | 10.7 | 54.6 |
| 7 | 683,037 | 10.2 | 69.4 | 10.7 | 47.7 | 11.3 | 52.0 | 11.2 | 65.8 |
| 8 | 683,036 | 10.2 | 79.6 | 12.5 | 60.2 | 12.7 | 64.7 | 11.2 | 77.0 |
| 9 | 683,037 | 10.2 | 89.8 | 15.1 | 75.3 | 15.2 | 79.8 | 11.1 | 88.2 |
| 10 | 683,036 | 10.2 | 100.0 | 24.7 | 100.0 | 20.2 | 100.0 | 11.8 | 100.0 |
| **Gini/concentration index** |  |  |  |  | 0.31 |  | 0.25 |  | 0.07 |
| **Kakwani index** |  |  |  |  |  |  | -0.06 |  | -0.24 |

Notes:

### Out-of-pocket costs in relation to equivalised household disposable income (progressivity)

Mean out-of-pocket cost as a percentage of equivalised household disposable income for both MBS and PBS decreased with income (Table 2). Percentages decreased from 1.16% in the poorest decile to 0.63% in the richest decile for MBS costs, and from 1.35% to 0.34% for PBS costs (Figure 1). The regressive trend was less pronounced for MBS, with percentage costs relatively stable between the 2^rd^ and 9^th^ deciles at 0.70-0.79% of income, but with percentages notably higher in the poorest decile and lower in the richest decile.

**Figure 1:**
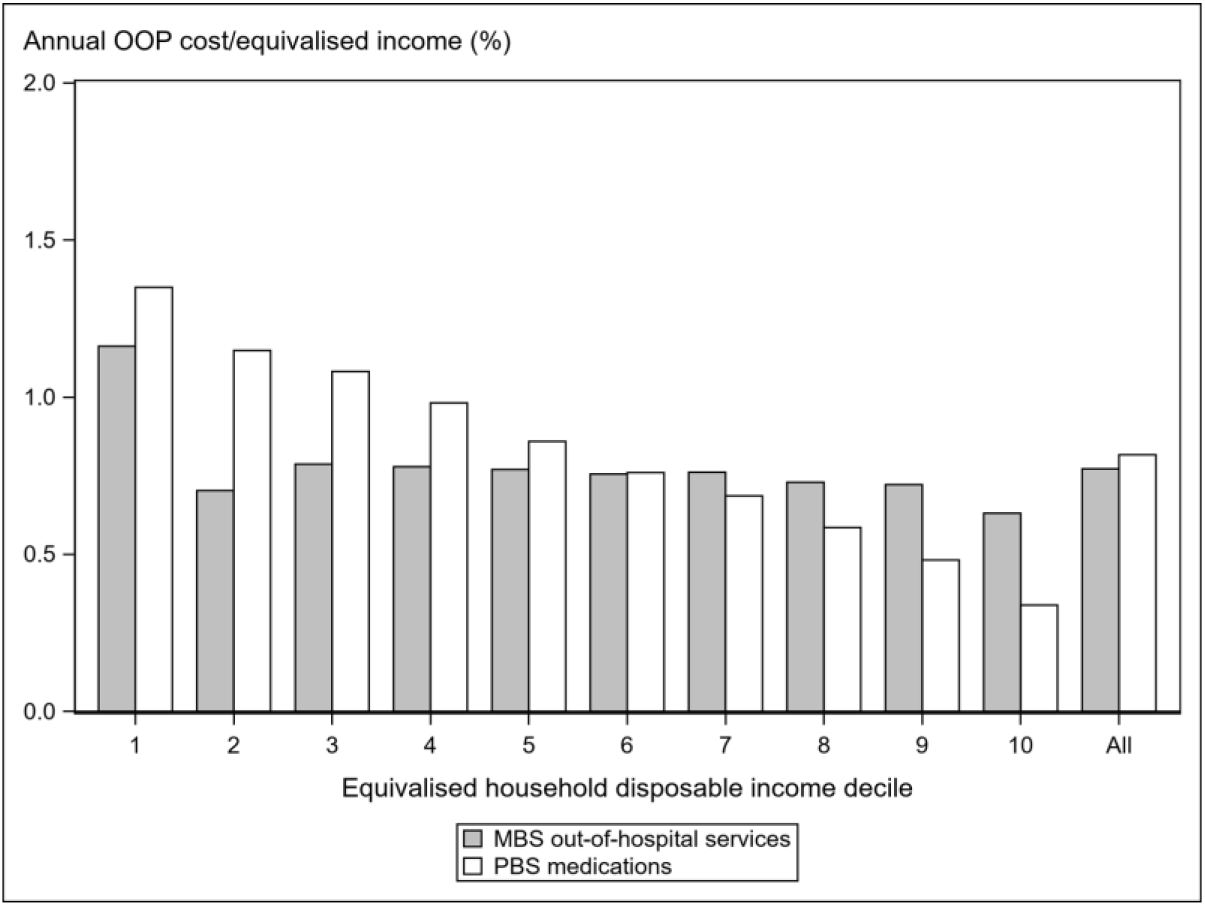
Annual out-of-pocket (OOP) costs as a percentage of equivalised household disposable income for Medicare-subsidised services, financial year 2017-18. Notes: 1. See Table 1 for sample sizes and Table 2 for total out-of-pocket costs and costs as a percentage of equivalised household disposable income.

**Figure 2:**
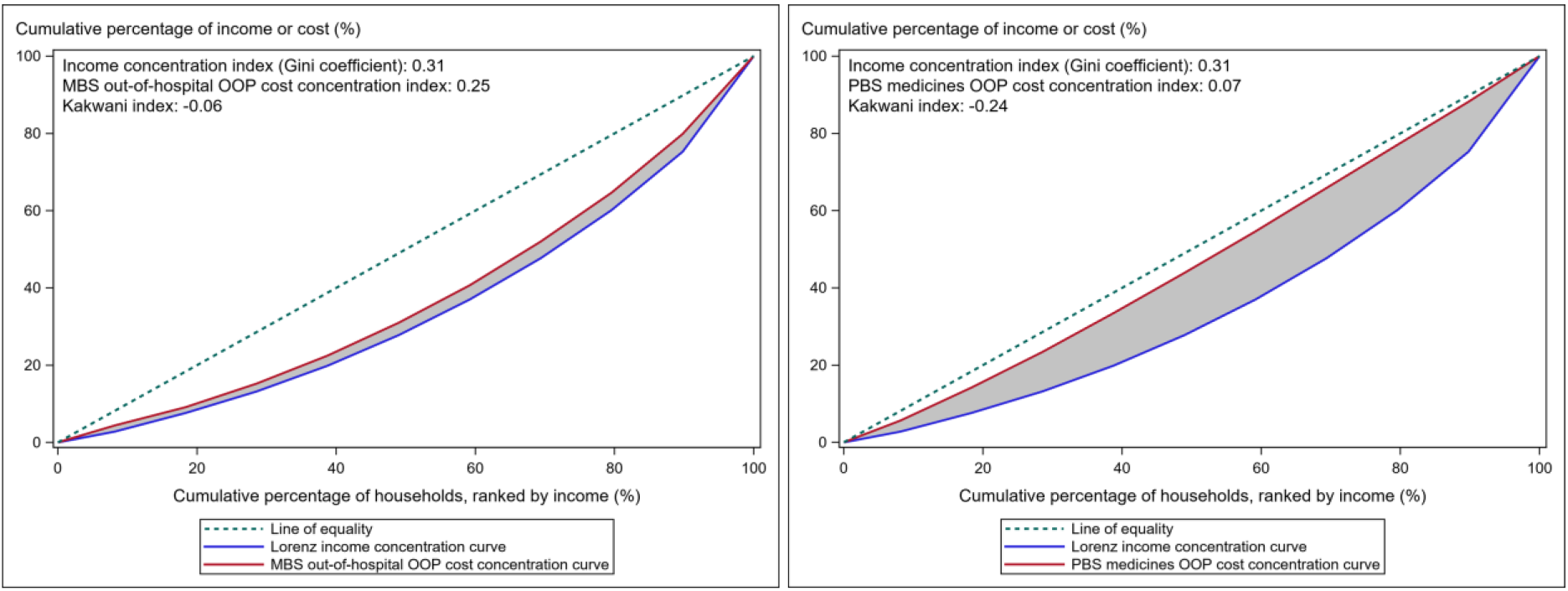
Concentration curves of equivalised household disposable income (financial year 2016-17) and out-of-pocket (OOP) costs for Medicare-subsidised services (financial year 2017-18), Australian households. Notes: 1. The concentration curve plots the cumulative percentage of households ranked from poorest to richest on the *x* axis, and the cumulative percentage of income (Lorenz curve) or cost on the *y* axis. 2 The extent to which the Lorenz curve lies outside (inside) the cost concentration curve represents the degree of regressivity (progressivity) of costs and is quantified by *K* (twice the area in between the Lorenz and cost curve). 3. See Table 3 for cumulative percentages of households, income and cost.

These results were reflected in the Kakwani indices, which showed that PBS out-of-pocket costs were more regressive than MBS out-of-pocket costs (*K*_*MBS*_ = −0.06, *K*_*PBS*_ = −0.24).

The Kakwani Index was largely insensitive to other methods of estimating disposable income (Supplementary Tables 2-3). Using income data from PIT or Census only, *K*_*MBS*_ was −0.07 and −0.08, and *K*_*PBS*_ −0.21 and −0.28 respectively. In supplementary analyses, we estimated percentage costs (Supplementary Tables 4-7) and the Kakwani Index by family type. The most regressive costs were observed among lone person households (*K*_*MBS*_ = −0.14, *K*_*PBS*_ = −0.33) followed by couple families without children (*K*_*MBS*_ = −0.12, *K*_*PBS*_ = −0.31), while single parent families had the least regressive costs (*K*_*MBS*_ = −0.06, *K*_*PBS*_ = −0.10). Couple families with children had similar cost distributions to the general population (*K*_*MBS*_ = −0.06, *K*_*PBS*_ = −0.20).

## DISCUSSION

Medicare guarantees all Australians access to a wide range of health services at low or no cost.^17^ In this study, we describe how closely out-of-pocket costs for some Medicare services relate to disposable income, shedding light on the financial burden of households according to their ability to pay. Understanding this distribution provides one measure of equity and an opportunity to reflect on the equity goals and affordability of Medicare. We report that in 2017-18, out-of-pocket costs for out-of-hospital MBS services were mildly regressive, while costs for PBS medicines were distinctly regressive. MBS out-of-pocket costs were approximately proportional to equivalised household disposable income at all levels except at the first and last deciles.

Consistent with known relationships between socioeconomic position and health, poorer households accessed more MBS and PBS services.^18^ Despite higher volumes of services among poorer households, out-of-pocket costs for MBS services were ‘pro-rich’ or concentrated among richer households. Our concentration index estimate (0.25) was very similar to those reported for those in the AusHEART survey (55 years and above) linked to MBS data (0.20-0.23 for out-of-hospital services over 2008-2012).^19^

In contrast, median out-of-pocket cost for PBS medicines approximately tracked the volume of supplies, which were generally lower in richer households. PBS costs were therefore considerably less pro-rich than MBS costs. Our estimate (0.07) was just slightly higher than that reported for those in the AusHEART survey (−0.001 in 2009), however that study did not contain PBS data for medicines under the maximum co-payment.^20^ Including drugs below the maximum co-payment, as we did in this study, is expected to contribute towards pro-richness. This is because richer households pay extra costs for these under co-payment drugs, as the maximum co-payment is set at a higher amount for non-concessional patients.

A pro-rich distribution of costs is not necessarily a progressive distribution; the former means that richer households paid a greater share of total cost, the latter depends on how absolute income itself is distributed. While out-of-pocket costs for MBS and PBS were both pro-rich, income was distributed pro-rich to an even greater extent—resulting in overall regressivity. We did not identify any studies that reported Kakwani indices for MBS and PBS costs exclusively. Although not directly comparable, Hajizadeh et al.^21^ reported a Kakwani index of −0.05 in 2009-10 for ‘direct payments’ (all health expenses including private health insurance (PHI) premiums) based on ABS Household Expenditure Surveys of self-reported data. We expect direct payments to be less regressive than the costs for Medicare services alone, as wealthier households are more likely to purchase PHI and the cost of non-Medicare services is likely to be higher in this group as they exercise their PHI policies.^22^

The reason that out-of-pocket costs for MBS services were only mildly regressive was possibly due to bulk-billing rather than safety net arrangements. It was not possible to identify households that were caught by the safety net with our data, however >90% of households in the bottom three deciles had costs below the lower concessional Extended Medicare Safety Net (EMSN) threshold, suggesting minimal impact of the EMSN on distributional equity (Supplementary Table 1). Poorer households were much more likely to be bulk-billed than richer households (Table 2). Providers of MBS services, unlike those for PBS medicines, are free to bulk-bill and set the fee charged at their discretion (and as such, determine the amount of out-of-pocket cost). Doctors also receive incentive payments to bulk-bill concession card holders.^9^ Previous studies have shown that bulk-billing is associated with lower income^23^ and that specialists tend to charge higher fees to higher income patients.^24^ Patients from lower income households may also self-select into bulk-billing practices, more so than patients from high income households.

In contrast, while concessional patients are entitled to a lower co-payment for PBS medicines, pharmacies do not practise further price discrimination based on income. The non-discretionary pricing of PBS medicines is likely to contribute to its regressivity. It was not possible to determine the impact of the PBS Safety Net on equity from our data, however 20-30% of households in the bottom three deciles had costs above the concessional PBS Safety Net threshold (Supplementary Table 1).

Among the strengths of our study was the inclusion of more than 80% of private households in the Census across the entire range of income distribution. The large sample size also allowed separate analysis of subgroups, including single-parent families. We used administrative data on out-of-pocket costs and income for the majority of our sample, avoiding error inherent in self-reported data.

PIT data enabled accurate calculation of post-tax income including for those at the very top of the income distribution. However, PIT data were not available for 27% of adults in our sample who did not complete a tax return. For these adults, we estimated income as the higher of Census income or Centrelink payments. We do not know which of these were more representative of disposable income in this group, nor the extent of under or over-estimation in either case. We conducted sensitivity analyses to avoid combining income concepts across datasets and these results did not change our conclusions.

PIT data do not contain information on income outside the tax system, for example, tax-exempt scholarships, inheritances and tax-free superannuation. Superannuation is unlikely to be a significant source of income for most individuals who complete a tax return and we assume the impact of other tax-exempt income was minimal. For retired individuals outside of the tax system, we relied on the Census to capture superannuation income as part of total income.^d^ It was not possible to verify the accuracy of this data. If the amount of superannuation was systematically under-reported, the extent of regressivity in costs may be over-estimated. Finally, we assumed that family composition as reported in the 2016 Census remained the same (apart from deaths, which we accounted for) at the time costs were incurred in 2017-18.

While we reported average trends, the data showed that households within any particular decile experienced a wide range in costs. This variation tended to increase with income. It should be noted that the costs considered in this study represent only a portion of all of out-of-pocket spending for healthcare. Individuals also incur costs for other allied services, medicines and dental treatment not subsided by Medicare, and for PHI premiums. Additionally, the extent of progressivity (or otherwise) in out-of-pocket payments does not reflect progressivity of the Medicare scheme as a whole. The latter is a weighted average of progressivity indices of all individual funding sources of Medicare. Medicare is primarily funded from taxation revenues and a levy on personal income, both of which have been shown to be progressive.^21, 25^

In summary, we report that out-of-pocket costs for out-of-hospital MBS services were slightly regressive but almost proportional to equivalised household disposable income, while out-of-pocket costs for PBS medicines were distinctly regressive. With the availability of linked data for a large majority of the Australian population, routine calculation of progressivity indices as performed in this study will allow monitoring of distributional equity over time and in response to policy. Measures to reduce inequity in out-of-pocket costs particularly for PBS medicines should be considered.

## Data Availability

All data produced in the present work are contained in the manuscript.

## Footnotes

a PIT and Census data are censored at $5,000,000 and $156,000 total income respectively.

b The tax-free threshold was $18,200 in 2016-17, but effectively $20,542 after taking into account the Low Income Tax Offset.

c Including some but not all tax-exempt incomes.

d For retirees who had PIT data, we added the amount of Census income in excess of PIT income to the disposable income estimate (see Eq. 2). For those without PIT data, we used Census income data alone. In both cases, we relied on people accurately including superannuation income as part of reported total income.

