## Supplementary material for "Progressivity of out-of-pocket costs under Australia’s universal health care system: a national linked data study"

**Supplementary Figure 1: Flow diagram of sample selection**

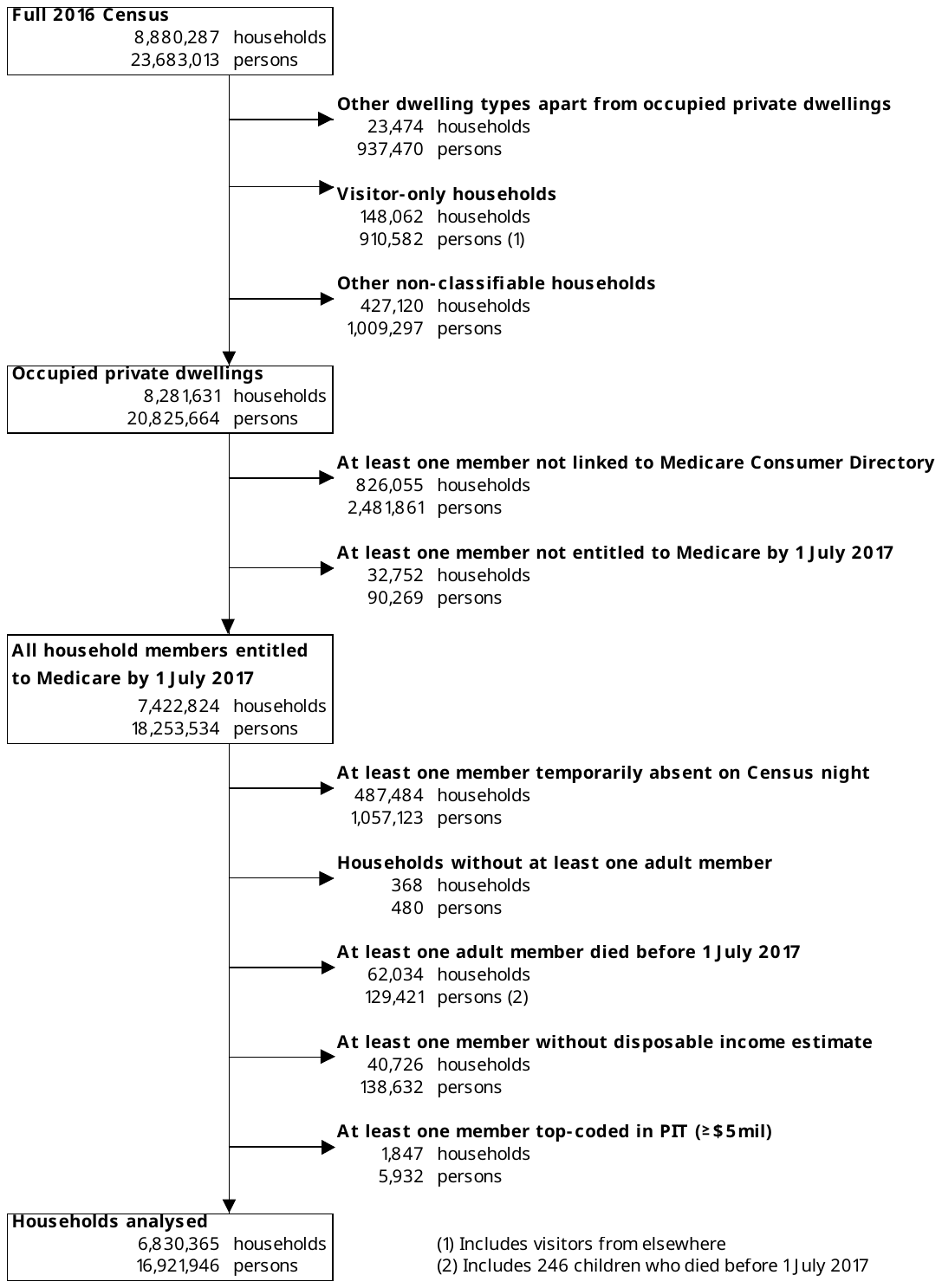

**Supplementary Figure 2. Percentage of adults whose total Centrelink payments were below, within or above their self-reported Census income, by Census (annual) income band, 2016-17.**

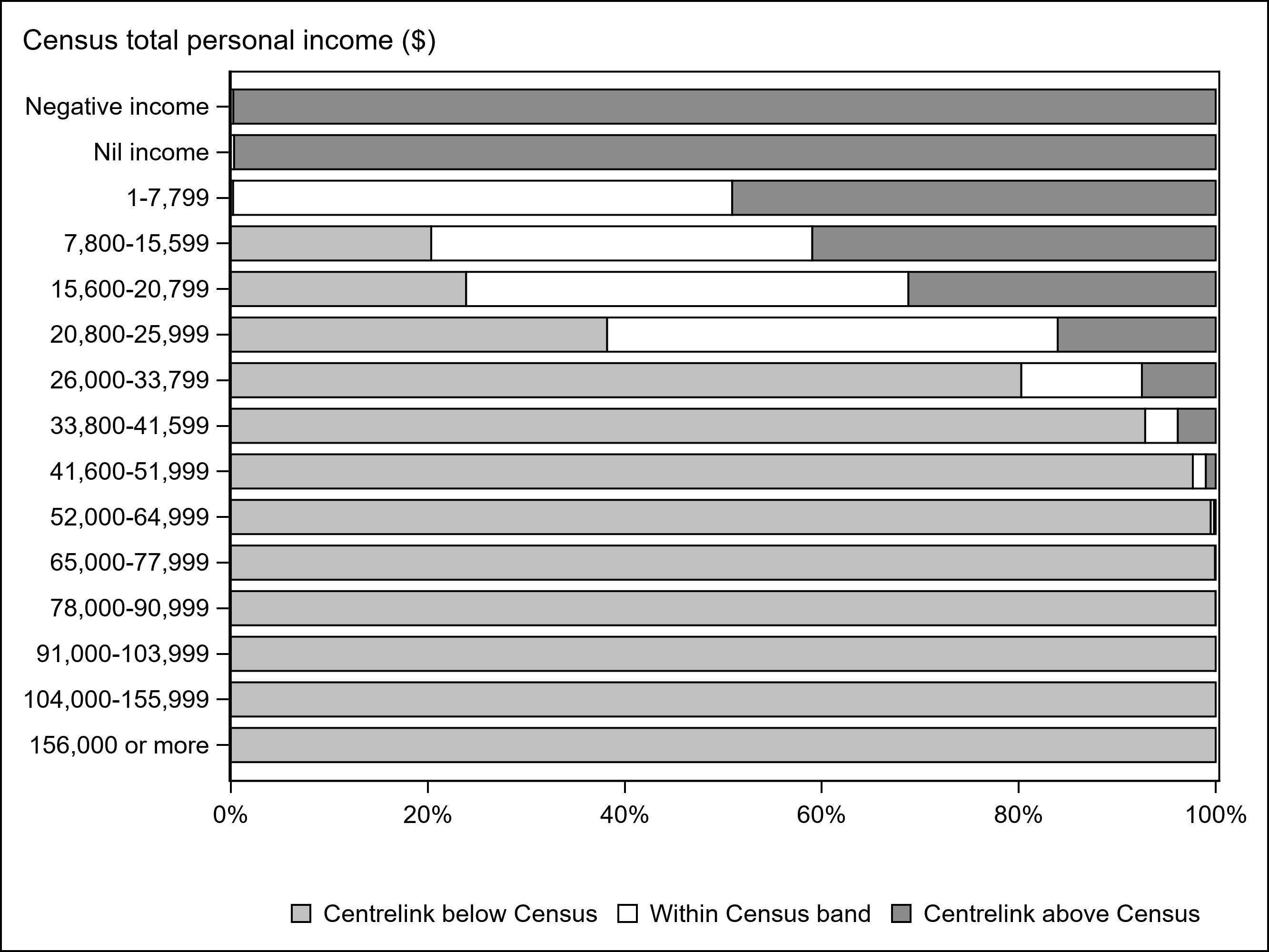

Notes:
1. Data included 5,287,526 adults in our study sample that had non-missing Census income data and at least one Centrelink benefit payment in 2016-17.

**Supplementary Table 1. Percentage of households where annual out-of-pocket costs were below the 2017 concessional thresholds for the Extended Medicare Safety Net or PBS Safety Net, Australian households, financial year 2017-18.**

| **Households** | | **MBS services** | **PBS medicines** |
| --- | --- | --- | --- |
| **Income decile** | **N** | **% below concessional Extended Medicare Safety Net** | **% below concessional PBS Safety Net** |
| 1 | 546,434 | 92.5 | 80.2 |
| 2 | 683,034 | 94.2 | 71.0 |
| 3 | 683,035 | 91.5 | 68.3 |
| 4 | 683,037 | 89.3 | 67.1 |
| 5 | 683,036 | 86.9 | 66.9 |
| 6 | 683,037 | 84.6 | 67.9 |
| 7 | 683,037 | 81.6 | 66.7 |
| 8 | 683,036 | 79.1 | 67.9 |
| 9 | 683,037 | 75.1 | 68.5 |
| 10 | 683,036 | 67.1 | 66.9 |

Notes:
1. See Table 1 for income cut-offs. Households in the first and second percentiles were excluded from the first income decile.
2. The concessional Extended Medicare Safety Net (EMSN) threshold in 2017 was $656.30.
3. The concessional PBS Safety Net threshold in 2017 was $378.
4. Our estimates accumulate out-of-pocket costs in the financial year, while the EMSN and PBS Safety Net are applied to the calendar year.

**Supplementary Table 2. Sample characteristics, service usage and annual out-of-pocket (OOP) costs for Medicare-subsidised services by income decile (income derived using PIT data only), Australian households, financial year 2017-18.**

| **Sample** | | | **MBS services** | | | | | **PBS medicines** | | | |
| --- | --- | --- | --- | --- | --- | --- | --- | --- | --- | --- | --- |
|  | **Mean** | **Mean** | **Mean** | **Proportion** | **Mean** | **Median** | **Mean** | **Mean** | **Mean** | **Median** | **Mean** |
| **Income decile** | **Household size** | **Age of adults** | **Number of services** | **Bulk-billed (%)** | **OOP  (SD) ($)** | **OOP (Q1-Q3) ($)** | **OOP/ equivalised income (%)** | **Number of supplies** | **OOP  (SD) ($)** | **OOP (Q1-Q3) ($)** | **OOP/ equivalised income (%)** |
| 1 | 2.6 | 42 | 31 | 0.89 | 206 (450) | 48 (0-245) | 1.21 | 17 | 181 (269) | 89 (25-228) | 1.06 |
| 2 | 2.7 | 42 | 32 | 0.86 | 254 (487) | 95 (0-322) | 0.90 | 17 | 231 (317) | 120 (34-301) | 0.82 |
| 3 | 2.6 | 42 | 31 | 0.85 | 281 (530) | 114 (0-355) | 0.78 | 17 | 255 (344) | 133 (38-336) | 0.71 |
| 4 | 2.6 | 41 | 31 | 0.84 | 314 (580) | 131 (0-395) | 0.74 | 16 | 270 (364) | 138 (39-355) | 0.64 |
| 5 | 2.6 | 41 | 32 | 0.83 | 354 (639) | 154 (0-443) | 0.73 | 16 | 286 (381) | 147 (41-378) | 0.59 |
| 6 | 2.6 | 40 | 32 | 0.82 | 388 (693) | 177 (20-484) | 0.72 | 16 | 293 (391) | 148 (42-388) | 0.54 |
| 7 | 2.6 | 40 | 32 | 0.81 | 431 (759) | 201 (33-533) | 0.71 | 16 | 300 (402) | 149 (42-396) | 0.49 |
| 8 | 2.5 | 41 | 32 | 0.79 | 485 (858) | 230 (43-587) | 0.70 | 16 | 301 (404) | 148 (40-399) | 0.44 |
| 9 | 2.5 | 41 | 32 | 0.77 | 574 (998) | 278 (66-684) | 0.71 | 16 | 302 (406) | 147 (39-403) | 0.37 |
| 10 | 2.4 | 43 | 31 | 0.72 | 737 (1,231) | 366 (96-876) | 0.61 | 16 | 313 (419) | 154 (39-422) | 0.26 |
| All | 2.6 | 41 | 32 | 0.82 | 406 (780) | 168 (0-488) | 0.77 | 16 | 275 (376) | 136 (38-360) | 0.58 |

Notes:
1. Households in the first and second percentiles were excluded from the first income decile. A total of 3,508,802 households were analysed.
2. Average proportions are 'mean of ratios' rather than 'ratio of means', that is, the mean of cost:income ratios across all households, rather than the ratio of the sum of cost to the sum of income.

**Supplementary Table 3. Sample characteristics, service usage and annual out-of-pocket (OOP) costs for Medicare-subsidised services by income decile (income derived using Census data only), Australian households, financial year 2017-18.**

| **Sample** | | | **MBS services** | | | | | **PBS medicines** | | | |
| --- | --- | --- | --- | --- | --- | --- | --- | --- | --- | --- | --- |
|  | **Mean** | **Mean** | **Mean** | **Proportion** | **Mean** | **Median** | **Mean** | **Mean** | **Mean** | **Median** | **Mean** |
| **Income decile** | **Household size** | **Age of adults** | **Number of services** | **Bulk-billed (%)** | **OOP  (SD) ($)** | **OOP (Q1-Q3) ($)** | **OOP/ equivalised income (%)** | **Number of supplies** | **OOP  (SD) ($)** | **OOP (Q1-Q3) ($)** | **OOP/ equivalised income (%)** |
| 1 | 2.2 | 53 | 41 | 0.93 | 151 (324) | 10 (0-177) | 1.18 | 39 | 239 (252) | 178 (51-369) | 1.82 |
| 2 | 1.8 | 60 | 37 | 0.92 | 149 (309) | 28 (0-182) | 0.69 | 39 | 243 (234) | 197 (70-363) | 1.11 |
| 3 | 2.4 | 58 | 49 | 0.91 | 224 (375) | 93 (0-307) | 0.84 | 53 | 312 (277) | 283 (103-433) | 1.18 |
| 4 | 2.6 | 50 | 43 | 0.89 | 255 (437) | 108 (0-345) | 0.75 | 38 | 315 (340) | 229 (73-438) | 0.93 |
| 5 | 2.8 | 48 | 43 | 0.87 | 313 (507) | 151 (0-425) | 0.74 | 35 | 342 (380) | 238 (80-458) | 0.81 |
| 6 | 2.9 | 44 | 41 | 0.85 | 364 (594) | 180 (20-480) | 0.70 | 29 | 360 (421) | 225 (70-485) | 0.70 |
| 7 | 2.6 | 44 | 36 | 0.83 | 388 (649) | 181 (10-502) | 0.62 | 24 | 342 (427) | 193 (53-462) | 0.55 |
| 8 | 2.7 | 43 | 36 | 0.80 | 470 (748) | 237 (43-605) | 0.63 | 22 | 349 (439) | 190 (55-471) | 0.47 |
| 9 | 2.5 | 43 | 33 | 0.78 | 523 (868) | 258 (50-654) | 0.57 | 19 | 331 (433) | 169 (43-447) | 0.36 |
| 10 | 2.3 | 45 | 32 | 0.74 | 687 (1,153) | 338 (83-826) | 0.52 | 20 | 347 (457) | 171 (41-470) | 0.27 |
| All | 2.5 | 49 | 39 | 0.86 | 357 (673) | 144 (0-444) | 0.71 | 32 | 320 (379) | 207 (63-431) | 0.80 |

Notes:
1. Households in the first and second percentiles were excluded from the first income decile. A total of 6,469,106 households were analysed.
2. Average proportions are 'mean of ratios' rather than 'ratio of means', that is, the mean of cost:income ratios across all households, rather than the ratio of the sum of cost to the sum of income.

**Supplementary Table 4. Sample characteristics, service usage and annual out-of-pocket (OOP) costs for Medicare-subsidised services by income decile, Australian couple families without children, financial year 2017-18.**

| **Sample** | | | **MBS services** | | | | | **PBS medicines** | | | |
| --- | --- | --- | --- | --- | --- | --- | --- | --- | --- | --- | --- |
|  | **Mean** | **Mean** | **Mean** | **Proportion** | **Mean** | **Median** | **Mean** | **Mean** | **Mean** | **Median** | **Mean** |
| **Income decile** | **Household size** | **Age of adults** | **Number of services** | **Bulk-billed (%)** | **OOP  (SD) ($)** | **OOP (Q1-Q3) ($)** | **OOP/ equivalised income (%)** | **Number of supplies** | **OOP  (SD) ($)** | **OOP (Q1-Q3) ($)** | **OOP/ equivalised income (%)** |
| 1 | 2.0 | 63 | 51 | 0.88 | 311 (482) | 166 (7-431) | 1.92 | 54 | 375 (338) | 337 (159-470) | 2.29 |
| 2 | 2.0 | 71 | 64 | 0.91 | 273 (374) | 158 (20-388) | 1.15 | 79 | 382 (217) | 389 (271-464) | 1.61 |
| 3 | 2.0 | 67 | 63 | 0.90 | 286 (430) | 153 (0-403) | 1.08 | 78 | 396 (284) | 389 (239-478) | 1.50 |
| 4 | 2.1 | 63 | 55 | 0.87 | 344 (487) | 205 (41-481) | 1.07 | 62 | 408 (349) | 375 (193-492) | 1.27 |
| 5 | 2.0 | 61 | 52 | 0.86 | 377 (532) | 228 (50-525) | 0.98 | 57 | 421 (387) | 373 (173-504) | 1.10 |
| 6 | 2.1 | 54 | 43 | 0.83 | 410 (644) | 224 (46-545) | 0.92 | 40 | 439 (466) | 325 (101-566) | 0.99 |
| 7 | 2.1 | 52 | 42 | 0.83 | 417 (683) | 220 (43-540) | 0.81 | 38 | 422 (466) | 301 (85-542) | 0.82 |
| 8 | 2.1 | 47 | 37 | 0.81 | 467 (790) | 235 (48-579) | 0.77 | 28 | 405 (487) | 232 (63-547) | 0.67 |
| 9 | 2.1 | 46 | 35 | 0.78 | 571 (998) | 280 (76-668) | 0.78 | 24 | 381 (479) | 199 (53-517) | 0.52 |
| 10 | 2.0 | 49 | 36 | 0.73 | 803 (1,317) | 410 (132-927) | 0.72 | 25 | 426 (525) | 224 (56-589) | 0.38 |
| All | 2.0 | 56 | 46 | 0.84 | 457 (811) | 231 (49-562) | 0.96 | 46 | 407 (425) | 330 (107-504) | 1.00 |

Notes:
1. Income cut-offs used were as derived for the whole population (see Table 1), therefore each income decile may not have 10% of total households in this sample.
2. Households in the first and second percentiles were excluded from the first income decile. A total of 1,807,688 households were analysed.
3. Only households with one family were included in this analysis.

**Supplementary Table 5. Sample characteristics, service usage and annual out-of-pocket (OOP) costs for Medicare-subsidised services by income decile, Australian couple families with children, financial year 2017-18.**

| **Sample** | | | **MBS services** | | | | | **PBS medicines** | | | |
| --- | --- | --- | --- | --- | --- | --- | --- | --- | --- | --- | --- |
|  | **Mean** | **Mean** | **Mean** | **Proportion** | **Mean** | **Median** | **Mean** | **Mean** | **Mean** | **Median** | **Mean** |
| **Income decile** | **Household size** | **Age of adults** | **Number of services** | **Bulk-billed (%)** | **OOP  (SD) ($)** | **OOP (Q1-Q3) ($)** | **OOP/ equivalised income (%)** | **Number of supplies** | **OOP  (SD) ($)** | **OOP (Q1-Q3) ($)** | **OOP/ equivalised income (%)** |
| 1 | 4.3 | 38 | 46 | 0.91 | 237 (452) | 80 (0-297) | 1.44 | 24 | 232 (291) | 135 (51-309) | 1.39 |
| 2 | 4.3 | 38 | 48 | 0.91 | 245 (467) | 91 (0-312) | 1.06 | 27 | 261 (309) | 160 (61-358) | 1.13 |
| 3 | 4.2 | 38 | 48 | 0.91 | 264 (469) | 111 (0-346) | 0.97 | 29 | 288 (329) | 180 (70-398) | 1.06 |
| 4 | 4.1 | 39 | 48 | 0.89 | 306 (505) | 146 (0-406) | 0.94 | 29 | 320 (359) | 202 (78-442) | 0.98 |
| 5 | 4.0 | 39 | 47 | 0.87 | 358 (567) | 184 (25-473) | 0.94 | 27 | 335 (380) | 210 (82-455) | 0.88 |
| 6 | 3.9 | 39 | 46 | 0.85 | 418 (636) | 227 (46-547) | 0.94 | 25 | 351 (400) | 219 (84-474) | 0.79 |
| 7 | 3.9 | 39 | 45 | 0.83 | 487 (706) | 277 (76-634) | 0.94 | 24 | 367 (417) | 225 (88-494) | 0.71 |
| 8 | 3.8 | 39 | 45 | 0.81 | 563 (781) | 336 (107-733) | 0.94 | 24 | 384 (437) | 235 (90-518) | 0.64 |
| 9 | 3.8 | 40 | 44 | 0.78 | 660 (878) | 409 (146-854) | 0.91 | 23 | 399 (451) | 244 (92-541) | 0.55 |
| 10 | 3.8 | 41 | 43 | 0.73 | 850 (1,072) | 546 (206-1,107) | 0.77 | 22 | 396 (453) | 240 (88-538) | 0.37 |
| All | 4.0 | 39 | 46 | 0.84 | 472 (735) | 245 (45-610) | 0.96 | 25 | 347 (402) | 211 (81-467) | 0.79 |

Notes:
1. Income cut-offs used were as derived for the whole population (see Table 1), therefore each income decile may not have 10% of total households in this sample.
2. Households in the first and second percentiles were excluded from the first income decile. A total of 2,058,036 households were analysed.
3. Only households with one family were included in this analysis.

**Supplementary Table 6. Sample characteristics, service usage and annual out-of-pocket (OOP) costs for Medicare-subsidised services by income decile, Australian one parent families, financial year 2017-18.**

| **Sample** | | | **MBS services** | | | | | **PBS medicines** | | | |
| --- | --- | --- | --- | --- | --- | --- | --- | --- | --- | --- | --- |
|  | **Mean** | **Mean** | **Mean** | **Proportion** | **Mean** | **Median** | **Mean** | **Mean** | **Mean** | **Median** | **Mean** |
| **Income decile** | **Household size** | **Age of adults** | **Number of services** | **Bulk-billed (%)** | **OOP  (SD) ($)** | **OOP (Q1-Q3) ($)** | **OOP/ equivalised income (%)** | **Number of supplies** | **OOP  (SD) ($)** | **OOP (Q1-Q3) ($)** | **OOP/ equivalised income (%)** |
| 1 | 3.0 | 36 | 35 | 0.93 | 142 (342) | 0 (0-157) | 0.89 | 19 | 146 (198) | 82 (26-185) | 0.90 |
| 2 | 3.0 | 36 | 36 | 0.93 | 133 (341) | 0 (0-143) | 0.58 | 22 | 157 (208) | 87 (31-204) | 0.69 |
| 3 | 2.8 | 38 | 39 | 0.93 | 152 (344) | 21 (0-177) | 0.56 | 27 | 199 (244) | 115 (38-277) | 0.74 |
| 4 | 2.7 | 44 | 44 | 0.92 | 186 (367) | 52 (0-230) | 0.57 | 40 | 290 (294) | 203 (70-429) | 0.89 |
| 5 | 2.7 | 43 | 43 | 0.91 | 217 (403) | 77 (0-277) | 0.57 | 38 | 312 (328) | 216 (76-446) | 0.82 |
| 6 | 2.6 | 42 | 41 | 0.89 | 256 (453) | 108 (0-333) | 0.58 | 33 | 329 (358) | 221 (79-457) | 0.74 |
| 7 | 2.6 | 43 | 40 | 0.87 | 298 (508) | 136 (0-390) | 0.58 | 31 | 346 (379) | 229 (82-479) | 0.67 |
| 8 | 2.5 | 43 | 39 | 0.86 | 342 (568) | 166 (12-445) | 0.57 | 30 | 358 (396) | 235 (83-491) | 0.60 |
| 9 | 2.5 | 43 | 38 | 0.84 | 390 (628) | 202 (37-515) | 0.54 | 27 | 361 (406) | 232 (79-495) | 0.50 |
| 10 | 2.4 | 44 | 36 | 0.80 | 503 (799) | 263 (55-661) | 0.47 | 26 | 366 (436) | 222 (71-495) | 0.35 |
| All | 2.7 | 41 | 39 | 0.90 | 220 (447) | 65 (0-271) | 0.62 | 29 | 262 (318) | 153 (51-367) | 0.75 |

Notes:
1. Income cut-offs used were as derived for the whole population (see Table 1), therefore each income decile may not have 10% of total households in this sample.
2. Households in the first and second percentiles were excluded from the first income decile. A total of 666,376 households were analysed.
3. Only households with one family were included in this analysis.

**Supplementary Table 7. Sample characteristics, service usage and annual out-of-pocket (OOP) costs for Medicare-subsidised services by income decile, Australian lone person households, financial year 2017-18.**

| **Sample** | | | **MBS services** | | | | | **PBS medicines** | | | |
| --- | --- | --- | --- | --- | --- | --- | --- | --- | --- | --- | --- |
|  | **Mean** | **Mean** | **Mean** | **Proportion** | **Mean** | **Median** | **Mean** | **Mean** | **Mean** | **Median** | **Mean** |
| **Income decile** | **Household size** | **Age of adults** | **Number of services** | **Bulk-billed (%)** | **OOP  (SD) ($)** | **OOP (Q1-Q3) ($)** | **OOP/ equivalised income (%)** | **Number of supplies** | **OOP  (SD) ($)** | **OOP (Q1-Q3) ($)** | **OOP/ equivalised income (%)** |
| 1 | 1.0 | 57 | 22 | 0.90 | 115 (276) | 0 (0-124) | 0.71 | 23 | 172 (219) | 102 (13-261) | 1.06 |
| 2 | 1.0 | 71 | 32 | 0.92 | 114 (235) | 0 (0-139) | 0.49 | 43 | 240 (179) | 229 (95-368) | 1.04 |
| 3 | 1.0 | 61 | 28 | 0.92 | 113 (257) | 0 (0-127) | 0.42 | 35 | 217 (210) | 178 (45-343) | 0.81 |
| 4 | 1.0 | 63 | 24 | 0.88 | 145 (304) | 28 (0-178) | 0.46 | 30 | 219 (234) | 169 (38-337) | 0.71 |
| 5 | 1.0 | 57 | 21 | 0.86 | 163 (333) | 38 (0-200) | 0.43 | 22 | 209 (271) | 124 (17-311) | 0.55 |
| 6 | 1.0 | 53 | 19 | 0.84 | 172 (377) | 39 (0-206) | 0.38 | 18 | 206 (290) | 98 (13-299) | 0.46 |
| 7 | 1.0 | 46 | 16 | 0.80 | 194 (455) | 45 (0-220) | 0.38 | 11 | 195 (314) | 62 (0-252) | 0.38 |
| 8 | 1.0 | 50 | 17 | 0.80 | 216 (509) | 56 (0-249) | 0.36 | 15 | 204 (312) | 77 (0-283) | 0.34 |
| 9 | 1.0 | 48 | 15 | 0.76 | 252 (587) | 76 (0-290) | 0.35 | 12 | 197 (318) | 60 (0-254) | 0.27 |
| 10 | 1.0 | 52 | 17 | 0.75 | 308 (675) | 97 (0-363) | 0.27 | 14 | 233 (364) | 77 (0-312) | 0.21 |
| All | 1.0 | 58 | 23 | 0.88 | 165 (394) | 33 (0-187) | 0.44 | 26 | 214 (261) | 142 (22-325) | 0.67 |

Notes:
1. Income cut-offs used were as derived for the whole population (see Table 1), therefore each income decile may not have 10% of total households in this sample.
2. Households in the first and second percentiles were excluded from the first income decile. A total of 1,785,310 households were analysed.
3. Only households with one family were included in this analysis.
